# Etiology and Outcomes of Acute Flaccid Paralysis in Adults: A Study in Tertiary Care Center

**DOI:** 10.1101/2020.05.27.20104620

**Authors:** Mohit Gupta, Sarav Daid

## Abstract

**INTRODUCTION:** The World Health Organization defines acute flaccid paralysis syndrome as rapid onset of weakness of an individual’s extremities, often including weakness of the muscles of respiration and swallowing, progressing to maximum severity within 1–10 days. We carried out this observational study in a tertiary care center in North India with the aim of determining the etiology and outcome of Acute Flaccid Paralysis in adults.

**METHODS:** In this observational study, all cases diagnosed with Acute flaccid paralysis admitted to the tertiary care center from 1st January 2015 till 30th June 2016 were enrolled. Ninety-seven consecutive patients of age above 18 years presenting with weakness of duration less than two weeks were enrolled after obtaining informed consent and various investigations were studied.

**RESULTS:** The study analyzed data of 97 patients presenting with flaccid paralysis. They were analyzed for the demographic data, clinical features, investigations, electrophysiological data, and results of the treatment. The most common etiology of acute flaccid paralysis in this entire population was GBS, which was responsible for 50.5% of the cases, followed by the Hypokalemic paralysis (25.8%) followed by neuroparalytic snake bite (15.5%).In this study, 48% of patients with hypokalemic paralysis had a secondary cause for their condition. Primary hypokalemic periodic paralysis occurred in 52% of patients.

**CONCLUSIONS:** In this observational study, GBS was found to be the most common cause for Acute Flaccid Paralysis followed by Neuroparalytic snake bite. Respiratory paralysis was an important risk factor associated with mortality.

## INTRODUCTION

Acute flaccid paralysis (AFP) is a clinical syndrome characterized by rapid onset of weakness of lower motor neuron type, including weakness of the respiratory and pharyngeal muscles, progressing to maximum severity within several days to weeks. AFP is a complex clinical syndrome with a broad array of potential etiologies that vary remarkably with age[1] Patients presenting with acute onset of quadriparesis or paraparesis pose a unique challenge to the clinician. An accurate and early diagnosis is the key to a positive outcome. Various studies are available in the pediatric population regarding AFP prevalence and differential diagnoses, mainly as an offshoot of the global polio eradication initiative [2]. However, few such studies have been conducted among adults. We evaluated cases of acute lower motor neuron type of weakness and analyzed the underlying spectrum of clinical conditions. So, the aims and objectives include:

1. To determine the changing etiology of Acute Flaccid Paralysis.
2. To determine the outcome associated with each etiology.

## METHODS

This was an observational study. All cases diagnosed with Acute flaccid paralysis admitted to the Department of Medicine & its allied specialties in Dayanand Medical College and Hospital, Ludhiana, Punjab from 1st January 2015 till 30th June 2016 were enrolled for study. Ninety-seven consecutive patients of age above 18 years presenting with weakness of duration less than two weeks were enrolled after obtaining an informed consent.

Investigations documented included: -

- Hemogram
- RFT
- LFT
- ESR
- Calcium, Magnesium, Phosphorus
- Electrocardiogram
- Chest XRay
- Creatinine kinase, LDH
- Urine Myoglobin (if needed)
- Serum Myoglobin (if needed)
- Urine porphyrins (if needed)
- ANA, ANCA, HIV ELISA (if needed)
- MRI Spinal Cord (if needed)
- Arterial Blood Gas Analysis (If needed)
- NCV studies (if needed)
- EMG (if needed)
- T3, T4, TSH (if needed)
- Urine Electrolytes (if needed)

### Diagnosis of Acute Flaccid Paralysis was made according to the following criterion

#### Notification Criteria

##### Clinical Evidence

A person of any age with acute flaccid paralysis WHO defines AFP syndrome as “characterized by rapid onset of weakness of an individual’s extremities, often including weakness of the muscles of respiration and swallowing, progressing to maximum severity within 1-10 days. The term ‘flaccid’ indicates the absence of spasticity or other signs of disordered central nervous system (CNS) motor tracts such as hyperlexia, clonus, or extensor plantar responses” (World Health Organization 1993 WHO/MNH/EPI/93.3. Geneva)

### METHOD OF COLLECTION OF DATA

Inclusion criteria:

Any case of Acute flaccid paralysis admitted to the Department of Medicine & its allied specialties in Dayanand Medical College and Hospital, Ludhiana, Punjab who is: -

1. >18 years but <60
2. Paralysis is acute in onset (<2 weeks duration)
3. Flaccid nature (absence of clonus, extensor plantars, spasticity, scissoring, criss cross gait)
4. Paraparesis
5. Quadriparesis with or without respiratory or bulbar muscle involvement.

Exclusion criterion:

1. Duration (>2 weeks)
2. Traumatic
3. Hemiparesis
4. Moribund patient
5. Uncooperative patient

Duration of study:

All the patients admitted in the Department of Medicine and its allied specialties in Dayanand Medical College & Hospital till 30th june 2016 were included.

Outcome:

The study helped to know the changing etiology of acute flaccid paralysis over the past decade, and also the outcome associated with each etiology.

## RESULTS

Acute flaccid paralysis (AFP) is “characterized by rapid onset of weakness of an individual’s extremities, often including weakness of the muscles of respiration and swallowing, progressing to maximum severity within 1-10 days. The term ‘flaccid’ indicates the absence of spasticity or other signs of disordered central nervous system motor tracts such as hyperreflexia, clonus, or extensor plantar responses.”[3]

With poliomyelitis nearing its elimination in the world, the other causes of AFP in children and adults have become significant. Unlike other studies which are important from epidemiological point of view, because the WHO is running polio eradication campaign, this study was mainly conducted to know the clinical characteristics and differential diagnosis of individual causes of AFP, including distribution by age, gender, and time.

The prospective study analyzed data of 97 patients presenting with flaccid paralysis between January 2015 till June 2016. They were analyzed for the demographic data (age, sex), clinical features, investigations, electrophysiological data, and results of the treatment. Medical Research Council (MRC) sum score was used for valuing the muscle strength from 0 to 5 in proximal and distal muscles in upper and lower limbs bilaterally. Cranial nerve involvement was noted along with respiratory muscle weakness which was assessed by need of mechanical ventilation, oxygen administration, non-invasive ventilation and spirometer record. Sensory system and autonomic abnormalities were analyzed.

Maximum number of individuals were between the age of 20-40 (55.7%), 27.8% between the age of 30-40, 6.2% between the age of 17-20 and 10.3% above 60 years of age.

There was noted to be a male preponderance among patients with acute flaccid paralysis Males constituted 69/97 (71.1%) and females 28/97(28.9%) of total number of cases.

Table (1) shows the symptoms and their respective percentages with which the patients presented. Weakness of limbs (paraparesis with/without quadriparesis) was seen in 100% of patients. Diarrhea was present in 5.15% of patients, Respiratory distress in 38.14% and facial deviation/weakness and bulbar weakness in 10.31% and 2.06% respectively. Ocular symptoms in the form of ptosis/diplopia were present in 13 patients (13.4%). Sensory symptoms in the form of paresthesia/numbness/tingling were present in 12.37% patients and 2.06% patients had bowel/bladder complaints. Figure 1 shows the power at presentation among the patients. Power 3/5 at presentation was seen in 46/97 patients which included 24 patients of GBS, 13 patients of snake bite, 5 patients of hypokalemic paralysis, 2 patients of dermatomyositis and 1 patient each of porphyria and polymyositis. Power of 0/5 was seen in 8 patients of hypokalemic paralysis and 2 patients of GBS. Power of 1/5 was seen in 9 patients of GBS and 1 patient of porphyria. Power of 2/5 was seen in 8 patients of hypokalemic paralysis, 7 patients of GBS, 2 patients of polymyositis and 2 patients of snake bite.

**TABLE 1:** SYMPTOMS AT PRESENTATION.

| Chief complaints | Frequency | Percent |
| --- | --- | --- |
| **Weakness of limbs** | 97 | 100 |
| **Fever** | 5 | 5.15 |
| **Diarrhea** | 5 | 5.15 |
| **Respiratory distress** | 37 | 38.14 |
| **Facial weakness/deviation** | 10 | 10.31 |
| **Bulbar weakness** | 2 | 2.06 |
| **Ocular (Diplopia + Ptosis)** | 13 | 13.40 |
| **Sensory(Paresthesia/ Tingling/ Numbness)** | 12 | 12.37 |
| **Bladder/Bowel complaints** | 2 | 2.06 |

**FIG 1:**
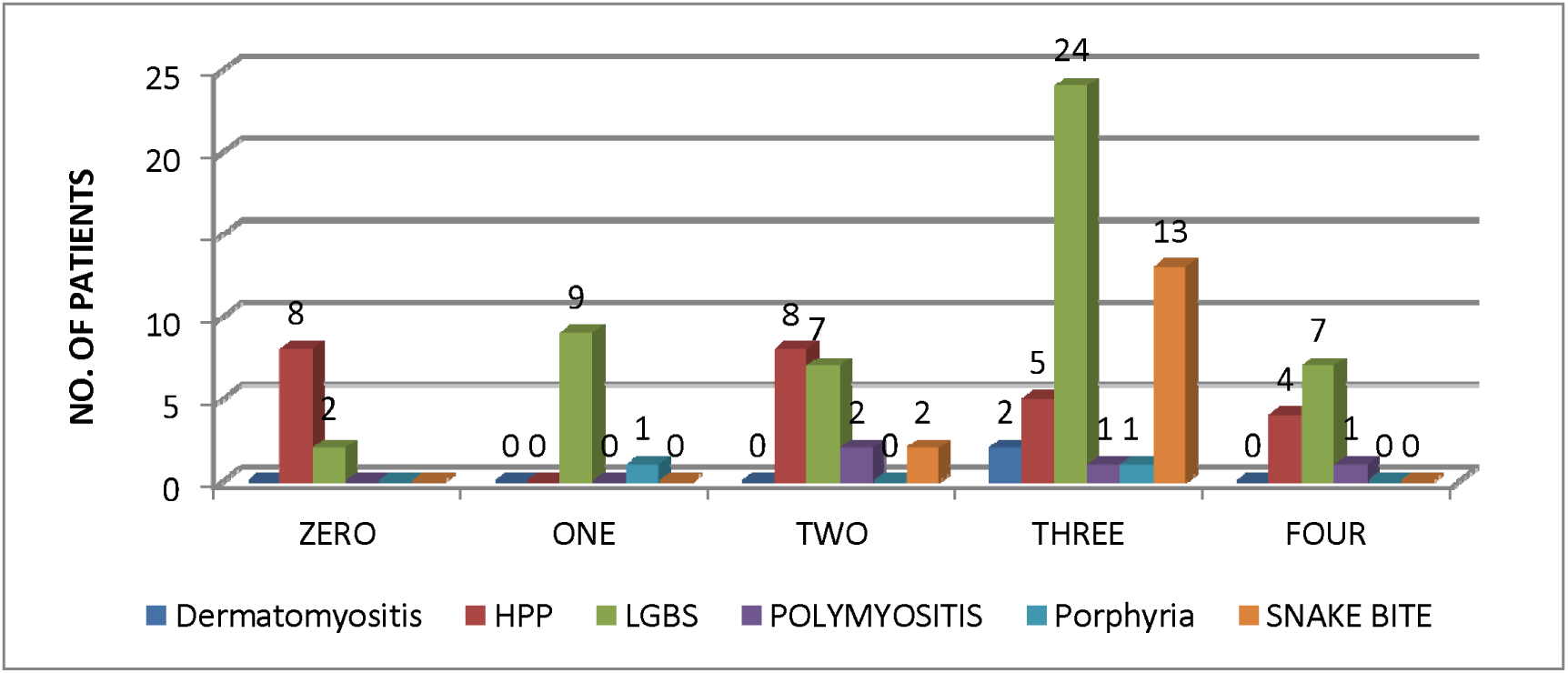
POWER AT PRESENTATION AND THEIR FINAL DIAGNOSIS.

In patients of GBS, 37/49 patients had the pattern of paralysis from lower to upper limb with 12/49 patients having the pattern of paralysis from upper limbs to lower limbs.

Positive ankle jerk at presentation was seen in 43/97(44.32%) patients. 93% patients of snake bite,76% patients of hypokalemic paralysis and 100% patients of polymyositis and dermatomyositis had positive ankle jerk at presentation.

Table (2) shows cranial nerve involvement in patients of different etiologies. Unilateral 7th nerve palsy seen in 5 patients of GBS and 1 patient of porphyria. Bilateral 7th nerve palsy was seen in 4patients of GBS. Bilateral 7th and bulbar palsy seen in 1 patient of GBS. 9th and 10th nerve palsy was seen in 1 patient of GBS and 4 patients of snake bite. External ophthalmoplegia seen in 1 patient each of GBS and snake bite.

**TABLE 2:** CRANIAL NERVE INVOLVEMENT AND DIAGNOSIS.

|  |  | Diagnosis 1 |  |  |  |  |  | Total |
| --- | --- | --- | --- | --- | --- | --- | --- | --- |
|  |  | Dermatomyositis | HP | LGBS | POLYMYOSITIS | Porphyria | SNAKE BITE |  |
| Cranial | 7th nerve palsy bilateral | 0 | 0 | 4 | 0 | 0 | 0 | 4 |
|  | 7th nerve palsy(rt+lt)+bulbar palsy | 0 | 0 | 1 | 0 | 0 | 0 | 1 |
|  | 9th+10th palsy | 0 | 0 | 1 | 0 | 0 | 4 | 5 |
|  | External | 0 | 0 | 1 | 0 | 0 | 1 | 2 |
| nerve | Ophthalmoplegia |  |  |  |  |  |  |  |
|  | Normal | 2 | 25 | 37 | 4 | 1 | 10 | 79 |
|  | unilateral 7th nerve palsy | 0 | 0 | 5 | 0 | 1 | 0 | 6 |
| Total |  | 2 | 25 | 49 | 4 | 2 | 15 | 97 |

Total of 15/97 patients had autonomic involvement in the form of postural hypotension, cardiac arrhythmias, resting tachycardia etc. This included 10 patients of GBS, 3 patients of snakebite and 2 patients of porphyria (Table 3).

**TABLE 3:** AUTONOMIC INVOLVEMENT IN PATIENTS OF DIFFERENT ETIOLOGIES.

| Diagnosis | AUTONOMIC INVOLVEMENT |  | Total |
| --- | --- | --- | --- |
|  | No | Yes |  |
| Dermatomyositis | 2 | 0 | 2 |
| Hypokalemic paralysis | 25 | 0 | 25 |
| LGBS | 39 | 10 | 49 |
| POLYMYOSITIS | 4 | 0 | 4 |
| Porphyria | 0 | 2 | 2 |
| SNAKE BITE | 12 | 3 | 15 |
| Total | 82 | 15 | 97 |

TABLE (4) shows the number of patients and their diagnosis who presented with respiratory distress at presentation. Total 37/97 patients had respiratory distress. This included 17 patients of GBS, 14 patients of snakebite, 4 patients of hypokalemic paralysis and 1 patient each of polymyositis and porphyria

**TABLE 4:** NUMBER OF PATIENTS AND THEIR DIAGNOSIS.

| Diagnosis | RESPIRATORY DISTRESS |  | Total |
| --- | --- | --- | --- |
|  | No | Yes |  |
| Dermatomyositis | 2 | 0 | 2 |
| Hypokalemic paralysis | 21 | 4 | 25 |
| LGBS | 32 | 17 | 49 |
| POLYMYOSITIS | 3 | 1 | 4 |
| Porphyria | 1 | 1 | 2 |
| SNAKE BITE | 1 | 14 | 15 |
| Total | 60 | 37 | 97 |

NCV findings in patients of GBS (axonal category) 13/25 had purely motor findings while 12 had sensorimotor. For NCV finding in demyelinating category of GBS, 14/24 patients had purely motor findings while 10 had sensorimotor.

The most common etiology of acute flaccid paralysis in this entire population was GBS, which was responsible for 50.5% of the cases, followed by the Hypokalemic paralysis (25.8%) followed by neuroparalytic snake bite (15.5%). Rest of the population consisted of representations of polymyositis, dermatomyositis and porphyria.

Figure (2) shows respiratory distress and requirement of NIV/VENTILATORY support and mean duration of ventilator support. 4 patients of hypokalemic paralysis had respiratory distress out of which 2 required ventilator support with mean duration of 4.5 days. 14 patients of snakebite had respiratory distress out which 12 required ventilator distress with mean duration of 4.5 days. 17 patients GBS had respiratory distress out of which 2 required NIV and 6 required ventilator support with mean duration of ventilator support 8.5 days. Solumedrol was received by 22 patients and IVIG by 29 patients of different etiologies.

**FIGURE 2:**
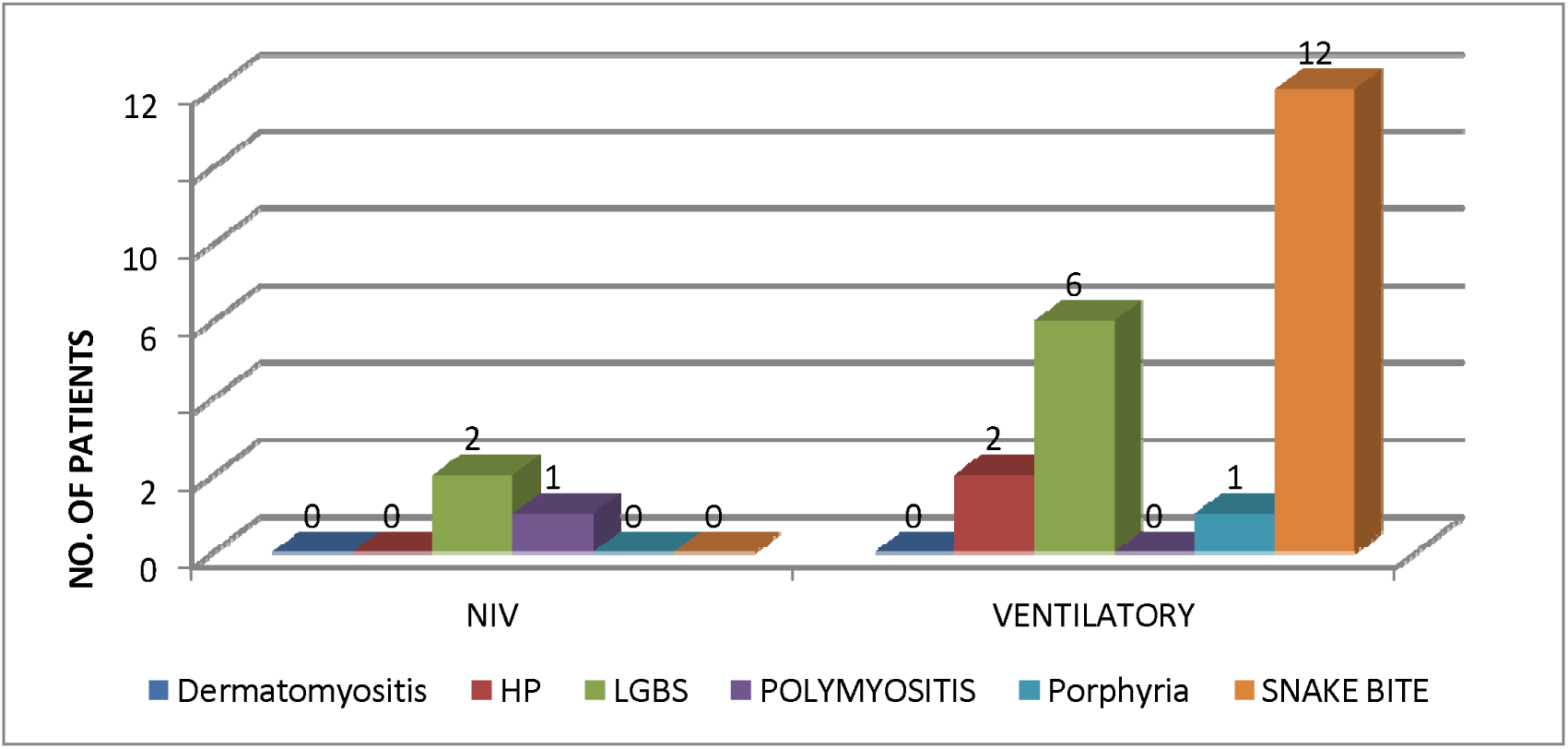
VENTILATORY REQUIREMENT AND DURATION IN DIFFERENT ETIOLOGIES.

The mean duration of hospital stay in patients of different etiologies was 7.8. Minimum duration of hospital stay was seen in patients of dermatomyositis of 4 and maximum in patients of GBS (9.27). Hospital stay in patients of hypokalemic paralysis and snake bite was 5.68 and 6.8 respectively.

Table (5) shows the complications which patient developed during hospital course. 22 out of total 97 patients developed complications. 5 patients of hypokalemic paralysis, 4 patients of GBS and 1 patient of dermatomyositis developed acute kidney injury. 1 patient of hypokalemic paralysis had AKI and pneumothorax. Bed sore developed in 4 patients of

**TABLE 5:** AKI: ACUTE KIDNEY INJURY.

| Diagnosis | COMPLICATIONS |  |  |  |  |  | Total |
| --- | --- | --- | --- | --- | --- | --- | --- |
|  | NO | YES(A<br>KI) | YES(AKI+PNE<br>UMOTHORAX | YES(ARRTH<br>MIAS) | YES(BE<br>D<br>SORE) | YES(P<br>NEUM<br>ONIA) |  |
|  |  |  | ) |  |  |  |  |
| Dermatomyositis | 1 | 1 | 0 | 0 | 0 | 0 | 2 |
| Hypokalemic paralysis | 19 | 5 | 1 | 0 | 0 | 0 | 25 |
| LGBS | 36 | 4 | 0 | 1 | 4 | 4 | 49 |
| POLYMYOSITIS | 3 | 0 | 0 | 0 | 1 | 0 | 4 |
| Porphyria | 2 | 0 | 0 | 0 | 0 | 0 | 2 |
| SNAKE BITE | 14 | 0 | 0 | 0 | 0 | 1 | 15 |
| Total | 75 | 10 | 1 | 1 | 5 | 5 | 97 |

GBS and 1 patient of polymyositis. Ventilator associated pneumonia developed in 4 patients of GBS and 1 patient of snakebite.

Figure (3) shows the outcome in patients of GBS who received either Solumedrol or IVIG. Among the patients who received Solumedrol 9 fully recovered, 4 partially recovered (at 2 months) 1 had a relapse and 2 patients expired. Among patients who received IVIG 21 patients fully recovered, 4 partially recovered (at 2 months) and 3 patients expired.

**FIGURE 3:**
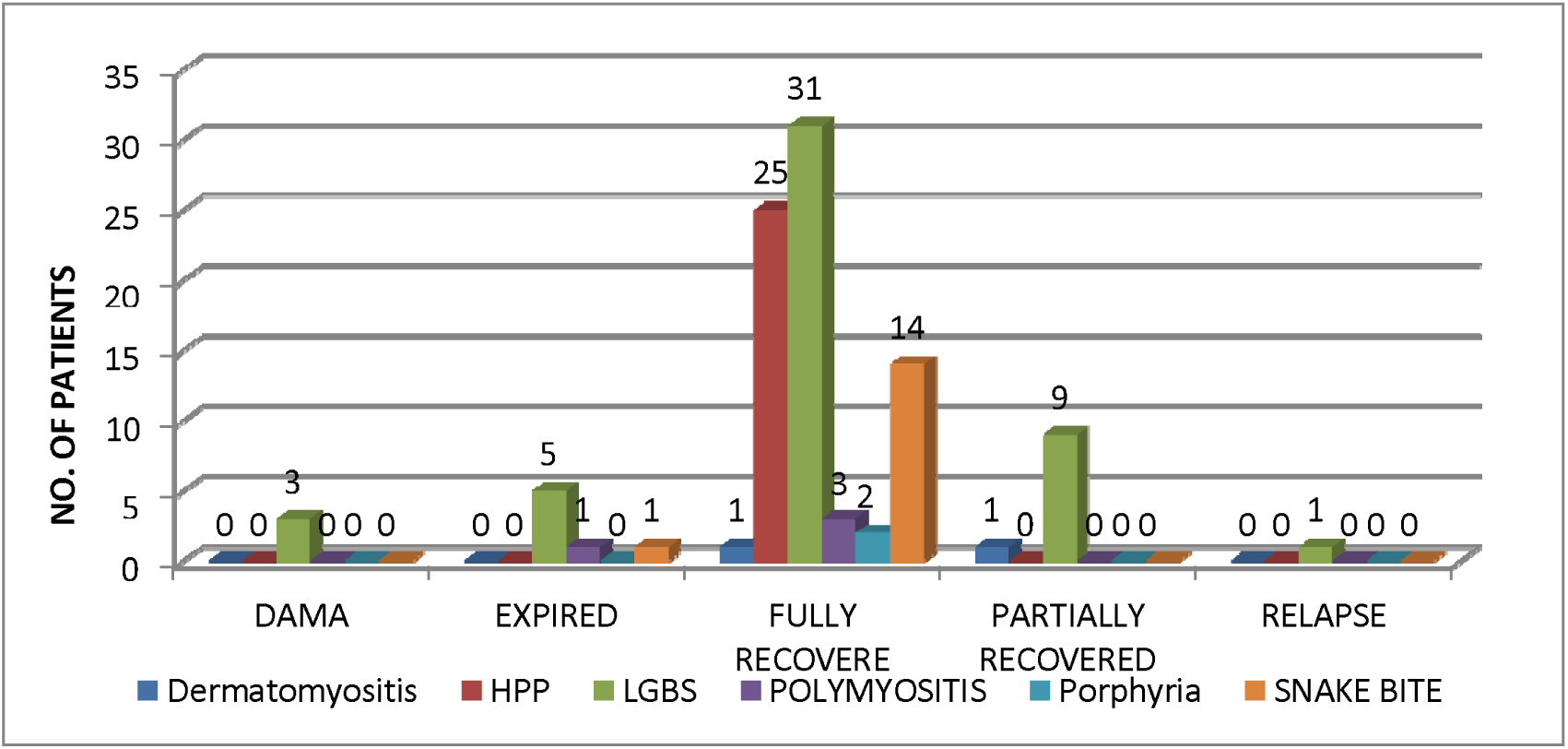
FINAL OUTCOME IN PATIENTS OF FLACCID PARALYSIS OF DIFFERENT ETIOLOGIES.

Figure 3 shows the final outcome in patients of flaccid paralysis of different etiologies. Among the 2 patients of dermatomyositis 1 fully recovered and 1 partially recovered (at 2 months follow up). All 25 patients of hypokalemic paralysis fully recovered without any residual disability or relapse. Among 49 patients of GBS 31 fully recovered, 9 partially recovered (at 2 months), 3 went dama, 5 expired and 1 had a relapse. Among 4 patients of polymyositis 3 fully recovered and 1 expired. 2 patients of porphyria had full recovered power at discharge. Among 15 patients of snake bite, 14 patients fully recovered and 1 expired.

## DISCUSSION

Historically, the predominant cause of AFP has been infection with poliovirus. The availability of an effective poliovirus vaccine led to a dramatic decline in poliovirus infections worldwide. Since the elimination of poliovirus from large parts of the world, Guillain-Barré syndrome (GBS) has become the most important clinical cause of AFP [4,5]. Various studies around the world have found prevalence of GBS among acute flaccid paralysis patients of 42-47%.[6] Higher prevalence has been reported from Honduras (72%)[7]. Studies around the world have identified envenomation, porphyria, hypokalemia, early acute transverse myelitis, rhabdomyolysis, botulism, and myasthenia gravis as other causes of acute flaccid paralysis. Neuroparalytic snakebite has been previously reported as a significant cause of acute flaccid paralysis in young rural men from Northern India. [2]

Our study shows a male preponderance among patients with acute flaccid paralysis Males constituted 69/97 (71.1%) of total number of cases. This is attributed to higher proportion of males in both snake envenomation and LGBS groups. Males are at a higher risk for snake envenomation due to occupational and recreational outdoor activities that predispose them to encounters with venomous snakes. The reasons for such a predilection are not clear. Male preponderance in GBS cases has been reported in various studies and our study reflects similar result. [8,9].

A preponderance of younger individuals was observed among the cases of acute flaccid paralysis .60.8% were less than 40years of age with 28.9% between the age group of 40-60. The results are similar to the study quoted by Marx et al.[10][3]

The most common etiology of acute flaccid paralysis in this entire population was GBS, which was responsible for 50.5% of the cases, followed by the Hypokalemic paralysis (25.8%) followed by neuroparalytic snake bite(15.5%).This is in contrast to the study done by Kaushik et al[102] where neuroparalytic snake envenomation was most common cause followed by Guillain Barre syndrome. Rest of the population consisted of representations of polymyositis, dermatomyositis and porphyria.

The proportion of patients remained consistent throughout the year except during monsoon when peak incidence upto 35% was seen. Our observation is similar to studies by Kaur et al [11]. Sharma et al. who reported a peak incidence between June–July and Sept–October [12] and Sriganesh K et al. in 2013, who reported increased occurrence of GBS during the months of June to August [13]. The study conducted at PGI by Kaushik et al [2] where 47% of acute flaccid paralysis patients were seen during the monsoon season.

Weakness that begins distally and progresses proximally with no or minimal sensory involvement could be a pointer towards GBS. In GBS 37/49 patients (75.5%) had ascending paralysis. Simultaneous upper and lower limb weakness was seen mainly in snake envenomation, which involves the neuromuscular junction. Similar pattern of weakness was also observed in hypokalemic paralysis, which causes myopathy.

Twelve patients had descending type of weakness, all of which were GBS. Similarly, patients who present with weakness in the cranial nerve distribution at onset are more likely to have a diagnosis of GBS. In the present series, patients of external ophthalmoplegia had a diagnosis either of neuroparalytic envenomation or GBS. Similarly, bilateral facial palsy has a high specificity and positive predictive value for the diagnosis of GBS.

Hence, in patients presenting with ascending paralysis, a diagnosis of GBS should be considered and in those with external ophthalmoplegia/ bilateral LMN facial palsy at presentation along with flaccid paralysis, diagnosis of GBS is more likely. LGBS and snake envenomation accounted for 100 % of patients with bulbar paralysis. Bulbar weakness was seen in 26.66% of neuroparalytic snakebite patients, which is lower than that reported in previous retrospective studies [14]. Cranial nerve involvement was seen in 24.4% cases of GBS with cranial nerve VII involved in most of the cases as against 50% in a study by Morris et al [15] to 21% in a study by Olive et al. [16] with CN VII most often affected.

Power was analyzed in accordance with guidelines given by Medical Research Council. 48.9% patients of GBS presented with power of 3/5 at presentation. Power of 0/5 at presentation was maximally seen in patients of hypokalemic paralysis (8/25), compared to 2/49 patients of GBS. However, majority of patients of hypokalemic paralysis had power 2/5 at presentation.

Positive ankle jerk at presentation was seen in 43/97(44.32%) of patients. Out of these 93% patients of snake bite,76% patients of hypokalemic paralysis and 100% patients of polymyositis and dermatomyositis had positive ankle jerk at presentation.

Autonomic involvement of the cardiovascular system was noted in 15.5% of the population, out of which 66.6% had LGBS, 13.3% had acute intermittent porphyria, and 20% had snake envenomation. The figure is lower compared to other studies. [17,18] Overall, 22.7% (22/97) cases had sensory involvement. GBS accounted for most cases with sensory involvement, with 24.7% belonging to demyelinating and 25.8% to axonal category on NCV studies.

Respiratory involvement was seen in 38.2% of our patients and 56.7% of them required mechanical ventilation. This is significantly higher than the respiratory involvement seen in similar case series published from other parts of world. [8,9]. This difference could be due to an overrepresentation of neuroparalytic snakebite patients who had severe respiratory involvement. 14/15 (93.33%) patients of snake bite had respiratory distress at presentation and 12/14(85.7%) required ventilatory support. The figure is higher compared to study by Sharma N et al where 75.6% required ventilator support [14]. Patients with axonal variants of GBS who have more severe illness may also have contributed to the higher mechanical ventilatory requirement. Similar to previous studies, we also found predominance of axonal involvement in our GBS patients with respiratory distress [19]. The higher percentage of respiratory involvement in the present study may also be a reflection of referral bias with more severe cases requiring mechanical ventilation being referred to our hospital. The mean duration of ventilatory requirement was greater in GBS (8.5 days) as compared to neuroparalytic snake bite (4.5 days) and hypokalemic paralysis (4.5 days) and others. Though the ventilatory requirement was greater in neuroparalytic snake bite compared to others (85.7%), possibly because of late presentation at tertiary care or due to greater degree of respiratory distress at presentation but the mean duration of ventilator stay was less.

In this study, 48% of patients with hypokalemic paralysis had a secondary cause for their condition. Another study by Garg RK et al shower similar results [20]. Primary hypokalemic periodic paralysis occurred in 52% of patients, out of which 84% were sporadic while 16% were familial. In this study, distal RTA was found in 8% of cases. An earlier study from Kashmir has reported 21 cases of hypokalemic paralysis secondary to distal RTA over a period of 8 years [21]. In the South Indian study, RTA was the cause in 13 out of 31 (42%) cases of HPP [22]. In the study by Maurya et a l [23] RTA was the cause of HPP in 4 out of 30 (13.3%) cases; 3 cases were of distal RTA and only 1 case had proximal RTA. Nine of our patients had distal RTA secondary to Sjogren’s syndrome which accounted to 36% of all cases.

Hypokalemic paralysis in Sjogren’s syndrome may precede the more classic clinical findings and serves as a clinical marker for this diagnosis [24].

In our study, we had a case of hypokalemic paralysis following dengue fever, without any similar episode of weakness in the past. The pure motor quadriparesis in this patient had dramatic recovery after potassium supplementation. The patient was also investigated to exclude other causes of hypokalemia, but no other secondary cause was detected. Hypokalemia, in association with infectious diseases, dengue fever in particular, have been reported and documented in up to 28% of serologically proven cases of dengue infection [25]. Indian studies have recently reported serologically confirmed cases of dengue infection with acute, pure motor, reversible quadriparesis due to hypokalemia [26]. Hypokalemia occurring in dengue fever can be secondary to re-distribution of potassium into the cells, transient renal tubular abnormalities leading to increased urinary potassium wasting and increased catecholamine levels secondary to infections and secondary insulin resistance leading to intracellular shift of potassium [26].

In our study, the serum potassium concentrations were lower in patients with secondary hypokalemic paralysis than in those with primary hypokalemic paralysis. Previous study showed no significant difference in potassium values between patients of primary and secondary hypokalemic paralysis [27]. But, in the study by Maurya and colleagues [23] the serum potassium concentrations were significantly lower in patients of secondary hypokalemic paralysis than in those with primary hypokalemic paralysis.

Outcomes in acute flaccid paralysis patients were skewed due to preponderance of snake envenomation, a potentially reversible condition. 78/97(80.4%) had hospital stay of less than 10days.Among 4 patients who had hospital stay of more than 20days, were diagnosed as case of GBS of which 3 completely recovered and 1 expired. Mean hospital stay duration was 7.8. Hospital stay duration was minimally seen in patients of hypokalemic paralysis and neuroparalytic snake bite, showing rapid reversibility of the condition, despite the fact that most of the patients of snake bite required ventilatory support.

22.6% (22/97) of all patients had one or other complication during hospital stay. Hospital-acquired pneumonia and bed sore were the most common complications (22%), seen exclusively in intubated patients and mostly in patients of GBS. This was followed by cardiac arrhythmia, predominantly in patients of LGBS and acute intermittent porphyria. Acute kidney injury was seen in a significant number of patients of hypokalemic paralysis. But Acute kidney injury was transient gradually normalizing during the hospital course. One of the patients of hypokalemic paralysis had AKI with associated pneumothorax, possibly center line associated (inserted outside). Exposure keratitis, upper respiratory tract infection, aspiration pneumonia were other complications witnessed.

Among treatment options 4 groups of patients (GBS, Porphyria, Polymyositis, Dermatomyositis) received either IVIG/SOLUMEDROL. Among the 4 patients of polymyositis, all of them received solumedrol with 3 of them completely recovered and 1 took discharge against medical advice and eventually expired after 3days because of respiratory distress. Among 2 patients of dermatomyositis one of them received solumedrol and completely recovered at discharge while the other who refused for any treatment options had partial recovery on follow up. Among the 2 patients of porphyria IVIG and SOLUMEDROL was received by one each and both of them had complete recovery on follow up, though the patient who received IVIG had respiratory distress and required ventilatory support for 5 days.

Among patients of GBS 14/49(28.5%) received solumedrol and 28/49(57.1%) received IVIG. One of the patients due to financial constraints initially received solumedrol but later IVIG due to non-improving status. Among patient who received solumedrol 7(43.7%) had respiratory distress at presentation out of which 2/7(28.5%) required ventilatory support. Out of these 2(12.5%) expired and 1(6.2%) had a relapse. In total 9 patients (56.2%) fully recovered and 4(25%) had partial recovery at 2 months. Among the patients who received IVIG 7/28(25%) had respiratory distress at presentation out of which 4/7(57.1%) required ventilatory support. Out of these 7 patients 4/28(14.2%) had complete recovery and 2/7(28.5%) expired. In total among the patients who received IVIG, 21/28(75%) had complete recovery, 4/28(14.2%) had partial recovery at 2months and 3/28(10.7%) expired and none of the patients had a relapse.

Prognosis of neuroparalytic snakebite patients was excellent if care was provided before development of complications. This can be explained by the fact that 14/15(93.3%) had respiratory distress at presentation and 12/14(85.7%) required ventilator support. 14/15(93.3%) had a complete recovery showing the transient nature of weakness.

In total 7 patients expired. 1 belonging each to snake bite and porphyria, which had respiratory distress and required ventilatory support but refused possibly due to financial constraints. Among the 5 patients of GBS which expired, 4 of them developed nosocomial pneumonia following non-invasive ventilation/ventilatory support and further went into sepsis and hemodynamic instability and 1 of them had history of coronary artery disease and developed arrythmias and further shock and expired.

In this study, recovery with potassium replacement therapy was seen in all cases. The secondary group needed longer time to recover compared to the patients with primary HPP. This delay in recovery can be attributed to a significantly negative total body potassium balance in patients with secondary hypokalemic paralysis. There was no mortality during the entire period of this study, thereby suggesting that a timely intervention can be lifesaving in this easily treatable but potentially fatal disease.

## SUMMARY AND CONCLUSION

This was an observational study. All cases diagnosed with Acute flaccid paralysis admitted to the Department of Medicine & its allied specialties in Dayanand Medical College and Hospital, Ludhiana, Punjab from 1st January 2015 till 30th June 2016 were enrolled for study. Ninety-seven consecutive patients of age above 18 years presenting with weakness of duration less than two weeks were enrolled after obtaining an informed consent.

The results were the following:

1. Our study shows a male preponderance among patients with acute flaccid paralysis Males constituted 69/97 (71.1%) of total number of cases. Male preponderance in GBS cases has been reported in various studies and our study reflects similar results.
2. A preponderance of younger individuals with 60.8% were less than 40years of age and 28.9% between the age group of 40-60.
3. The most common etiology of acute flaccid paralysis in this entire population was GBS, which was responsible for 50.5% of the cases, followed by the Hypokalemic paralysis (25.8%) followed by neuroparalytic snake bite (15.5%).
4. The proportion of patients remained consistent throughout the year except during monsoon when peak incidence upto 35% was seen.
5. Patients who present with weakness in the cranial nerve distribution at onset are more likely to have a diagnosis of GBS. In the present series, patients of external ophthalmoplegia had a diagnosis either of neuroparalytic envenomation or GBS.
6. Similarly, bilateral facial palsy has a high specificity and positive predictive value for the diagnosis of GBS. LGBS and snake envenomation accounted for 100 % of patients with bulbar paralysis. Cranial nerve involvement was seen in 24.4% cases of GBS with cranial nerve VII involved in most of the cases
7. Respiratory involvement was seen in 38.2% of our patients and 56.7% of them required mechanical ventilation. Patients with axonal variants of GBS who have more severe illness may also have contributed to the higher mechanical ventilatory requirement.
8. The mean duration of ventilatory requirement was greater in GBS (8.5 days) as compared to neuroparalytic snake bite (4.5 days) and hypokalemic paralysis (4.5 days) and others
9. In this study, 48% of patients with hypokalemic paralysis had a secondary cause for their condition. Primary hypokalemic periodic paralysis occurred in 52% of patients, out of which 84% were sporadic while 16% were familial.
10. 80.4% had hospital stay of less than 10days.
11. Hospital stay duration was minimally seen in patients of hypokalemic paralysis and neuroparalytic snake bite, showing rapid reversibility of the condition, despite the fact that most of the patients of snake bite required ventilatory support.
12. 22.6% of all patients had one or other complication during hospital stay. Hospital-acquired pneumonia and bed sore were the most common complications (22%), seen exclusively in intubated patients and mostly in patients of GBS
13. Prognosis of neuroparalytic snakebite patients and hypokalemic paralysis was excellent if care was provided before development of complications.
14. Respiratory paralysis is an important risk factor which defines outcome associated with flaccid paralysis of different etiologies.
15. 78.3% patients had full recovery at discharge or at 2 months follow up.10.3% had partial recovery at 2 months. 7.2% patients expired and respiratory paralysis was an important risk factor associated with mortality. 1.03% patients had a relapse of symptoms.

## Data Availability

There is full availability of all data referred to in the manuscript.

## IRB Approval

All relevant ethical guidelines have been followed, and all necessary IRB and ethics committee approvals have been obtained from Dayanand Medical College and Hospital’s ethics committee.

